# BNT162b2 COVID-19 VACCINATION AND ITS EFFECT ON BLOOD PRESSURE

**DOI:** 10.1101/2021.06.16.21258803

**Authors:** Toh Leong Tan, Sharifah Azura Salleh, Zuraidah Che Man, Michelle Hwee Peng Tan, Rashid Kader, Razman Jarmin

## Abstract

This study aims to explore the trends of systolic blood pressure (SBP), diastolic BP (DBP), mean arterial pressure (MAP) and pulse pressure (PP) before and 15-minutes after two doses of the BNT162b2 vaccine, which were administered 21 days apart. This vaccine safety active surveillance study was carried out on 15th-16th March (first dose) and 5th-6th April 2021 (second dose), in academic hospital.. Vaccinees above 18 years old, SBP, DBP, MAP and PP pre- and 15 minutes post-vaccination for both doses were analysed. Study outcomes were mean of BP, mean of BP changes, and BP trends measurements. A total of 287 vaccinees were included. A quarter (n=72) had decreased DBP ≥ 10mmHg (mean DBP deceased: 15mmHg, 95%CI: 14-17mmHg) after the first dose, and 12.5% post-second dose (mean DBP decreased: 13mmHg, 95%CI: 12-15mmHg). Post-first dose, 28.6% (n= 82) were found to have widened PP > 40mmHg. After the first dose, those who had elevated and decreased SBP ≥ 20mmHg were 5.2% and 4.9%, respectively. Eleven percent (n = 32) had decreased SBP ≥ 20mmHg post-second dose, nevertheless the psychology effect cannot be ruled out. The BNT162b2 vaccine was generally well tolerated. BP changes after vaccination emphasizes the need for monitoring.

## INTRODUCTION

In December 2019, Coronavirus disease 2019 (COVID-19) caused by the SARS-CoV-2 virus was detected, and subsequently declared as a global Pandemic by WHO on 11th March 2020^1^. This pandemic caused travel restrictions, economic recession, and loss of many lives. Given the pandemic urgency, new vaccines against the SARS-CoV-2 virus were developed and approved under the clause of Emergency Use Authorization on December 11, 2020^2^, including the BNT162b2 COVID-19 vaccine. Following implementation of vaccination, reports of adverse events for immunisation (AEFI) after the first dose of the BNT162b2 vaccine emerged^3^. These AEFIs are further classified into 5 subcategories^4^.In the US, notifications and reports of suspected severe allergic reactions and anaphylaxis following vaccination were captured in the Vaccine Adverse Event Reporting System (VAERS), the national passive surveillance (spontaneous reporting) system for adverse events after immunization^5^. Anaphylaxis is a life-threatening allergic reaction that occurs rarely after vaccination, with onset typically within minutes to hours.

Not all AEFIs have a causal relationship with the usage of the vaccine^6^. Because AEFI may affect a healthy individual after vaccination, prompt action needs to be carried out to manage it and to prevent the public from developing loss of confidence towards the vaccination program.

In our recent vaccination safety active surveillance (VSAS) on the BNT162b2 vaccine, we observed a few minor adverse events, including significant transient fluctuation on blood pressure (BP) post-vaccination as compared with pre-vaccination. Although the fluctuation of BP is not considered as part of the AEFI, these observations can significantly affect subjects with co-morbidities. This concern seems valid as there is a case series recently published on eight vaccinees who experience elevated high BP after receiving the vaccine^7^. This observation is a valid concern and warrants further investigation on the relationship between vaccine and BP levels. This information can be useful for the next phase of COVID - 19 vaccination program especially since it involves subjects with comorbidities.

Thus, in this study, we aim to report the database of our VSAS on the BP trend before and 15 minutes post -vaccination. We aim to explore the trends of systolic BP (SBP), diastolic BP (DBP), mean arterial pressure (MAP), and pulse pressure (PP) before and 15 minutes post first and second doses of the BNT162b2 vaccine. We hypothesize that the BNT162b2 vaccine associates with pre- and 15 minutes post-vaccination BP changes.

## METHODS

The active surveillance was carried out as part of COVID-19 vaccination program activities to monitor BNT162b2 vaccination safety among our hospital healthcare workers (HCW). Due to the retrospective nature of this study, it waived the need of informed consent. All vaccine-related data that were collected were kept in a secured location and only accessible to authorised personnel. The VSAS was set up by the Vaccination Committee of Hospital Canselor Tuanku Muhriz (HCTM) under the patronage of the Hospital Director, following reports of BP fluctuation and other adverse events post vaccination in other vaccination centres across the globe. This BNT162b2 COVID-19 VSAS program was carried out on 15th-16th March 2021 for the participants’ first dose and 5th-6th April 2021 for the second dose which was 21 days apart. The surveillance was done at the designated vaccination centre in HCTM, the teaching hospital of Universiti Kebangsaan Malaysia (UKM), located at Cheras, Kuala Lumpur, Malaysia. Subjects included in the surveillance were healthcare working (HCW) in HCTM, aged above 18 years, and received their doses as scheduled. Subjects with missing data on blood pressure were excluded. The study was approved by UKM Human Ethical Review Board (JEP-2021-302). The active surveillance was carried out as part of COVID-19 vaccination program activities to monitor BNT162b2 vaccination safety among our hospital healthcare workers (HCW). Due to the retrospective nature of this study, it waived the need of informed consent.

### DATA COLLECTION

The protocol for this surveillance was integrated into the existing vaccination protocol that consists of pre-vaccination health screening for premorbid conditions and history of allergic reactions. The vaccine recipients were asked if they had common comorbidities such as hypertension, diabetes, cardiovascular disease, stroke or transient ischaemic attack, asthma, and others. Allergy history was also surveyed. BP of vaccinees was measured pre- and 15 minutes post-vaccination. A standardised form was created to record age, gender, comorbidities, pre-, and post-vaccination BP measurements including any adverse reactions. During the observation period, any recipient noted to have high BP or AEFI was referred to the on-site staff clinic and the Emergency Department if deemed necessary.

### MEASURING OF BLOOD PRESSURE

The pre-vaccination BP measurement was recorded by trained personnel for each patient in the sitting position after a 5-min rest using a digital BP measuring machine (Connex ProBP 3400) with appropriately sized cuffs. Post-vaccination BP measurement was recorded 15 minutes after compulsory resting and observation. Two measurements were taken for all vaccinees. In those who recorded a BP difference of more than 10□mm□Hg for either systolic or diastolic BP, the third measurement would be repeated after a 5-min rest. The final recorded BP measurements were the mean of the second and third recorded measurements. Vaccinees with SBP ≥140mmHg had their heart rate assessed. If they were found to be tachycardic with a heart rate ≥100 beats/min, they were asked to rest for 5min, thereafter have their heart rate re-measured. Vaccinees were allowed to receive their injections if they are not tachycardic anymore after resting. Otherwise, their appointment would be rescheduled. For this group of vaccinees, the final recorded BP was the measurement taken after rest.

### THE HCTM VACCINATION PROTOCOL

Vaccine recipients were notified of their appointments via the *Mysejahtera* application which is an online phone application created by the Malaysian government as one of the means to monitor COVID-19 infection and track COVID-19 vaccination progress. On the date of vaccination appointment, the vaccinees confirmed their attendance by scanning the QR code of the vaccination site using *Mysejahtera*. A medical officer was on-site to answer any queries that may arise from the recipient. Once the written consent was obtained for vaccination, recipients were allocated to a designated area while awaiting their vaccination. Recipients would proceed to the vaccination booth when their turn arrives. In the vaccination booth, a staff nurse would check the identity of the recipient and explain each step of the vaccination process to the recipient. The vaccine was then administered, and the recipients were asked to rest in a resting area for a minimum of 15 minutes while being observed for any acute side effects. Once the observation was completed, the recipients were given some general advice about monitoring future side effects. A vaccine card was issued to the vaccinees upon completion of the first dose and electronic certificates were issued upon completion of the second dose.

### STATISTICAL ANALYSIS PLAN

#### Sample Size

This is a report on our hospital’s VSAS, where the sampling method was to actively include all consecutive subjects systematically from surveillance records. The sample size was calculated based on the interim data from our COVID-19 VSAS database. Sample size was calculated based on an effect size of 0.2, with 2-sided p-value less than 0.05. Total sample size required to achieve a statistically significant difference with a mean with power of 80% was 199 paired subjects.

### Data Analysis

Data analysis was carried out using IBM SPSS for WINDOWS (version 25.0). For descriptive analysis, participants’ characteristic and other categorical variables were expressed in frequency and percentages. Those who received both doses of vaccines were subjected to both-doses analysis. Those who received only the first dose, and those who received the first dose from both-doses group were subjected to a subgroup first-dose analysis. Analysis of participant age depended on the normal distribution and was presented as mean or median with corresponding standard deviation, SD, or interquartile range, and 95% confidence interval. Further analysis of the comparison between means was performed using Student’s T-test. Paired t-test was used for within-group comparison. All tests were considered significant at p< 0.05. Multiple imputation analysis was used to analyse missing data when it was more than 5%. A difference of SBP ≥ 20mmHg (SBP pre-vaccination – SBP post-vaccination) and DBP ≥ 10mmHg was considered significant. PP ≥ 40mmHg was classified as wide pulse pressure. Any difference of PP ≥ 10mmHg (PP pre-vaccination – PP post-vaccination) was considered a significant change. For BP trends, ‘elevated’ & ‘decreased’ trends were defined as an increased or decreased mean difference of ≥20mmHg for SBP, ≥10 mmHg for DBP and ≥10 mmHg for PP between pre- and 15-min post-vaccination, respectively. Normal range BP was recorded as “no change”.

## RESULTS

### STUDY POPULATION

A total of 443 consecutive vaccinees presented on 15^th^-16^th^ March 2021 for their 1^st^ dose of vaccine. They were included in the first-dose analysis. After 21 days apart, on 5^th^-6^th^ April 2021, from the same cohort, only 288 of them completed their second doses - these were included in the both-doses analysis. Given the cohort who are mainly healthcare workers, 143 vaccinees rescheduled their second dose to a date after 6th April 2021 due to logistic reasons. The logistic reasons include unavoidable working shift mismatch, on leave, or falling sick on the proposed vaccine schedule date. During the data analysis, we exclude one subject from both-dose analysis due to missing BP values, which contribute to 0.3% of missing data. There were 12 subjects (2.8%) with missing BP data in the first-dose analysis. The missing data were considered completely missing in random which required no further imputation analysis. The age for the study group ranged from 21 to 60 years old. The demography of the study population for both-doses analysis is shown in **Table 1**. The study inclusion flow is shown in **Figure S1**. A total of eight participants (2.8%) with hypertension were included in this surveillance. Details of their report is shown in Supplementary file **Table S1 <u>(Supplemental Data)</u>**. There were no severe adverse events from the COVID-19 vaccination or death during this surveillance period.

**Table 1:** Demographic and Clinical Characteristics of Participants who received Both Doses of BNT162b2 Vaccine.

| DEMOGRAPHIC CHARACTERISTICS | NUMBER OF VACCINEES (N=287) |
| --- | --- |
| Age, years, median (IQR) | 33 (31-39) |
| Age group, years, n (%) |  |
| 21-29 | 58 (20.2) |
| 30- 39 | 165 (57.5) |
| 40-49 | 54 (18.8) |
| 50-60 | 10 (3.5) |
| Gender, n (%) |  |
| Male | 76 (26.5) |
| Female | 211 (73.5) |
| Comorbidities, n (%) | 45 (15.7) |
| Asthma | 9 (3.1) |
| HTN | 8 (2.8) |
| Diabetic Mellitus | 7 (2.4) |
| IHD | 3 (1.0) |
| Allergy | 2 (0.7) |
| Renal Disease | 1 (0.3) |
| Others | 15 (5.2) |

### TRENDS AND MEANS OF SBP, DBP, AND PP CHANGES BETWEEN PRE-AND 15-MINUTE POST-BNT162B2 VACCINATION IN BOTH-DOSES ANALYSIS

We analysed the trend of BP changes for those who completed both doses of vaccines. The results are shown in **Table 2**. Percentage of vaccinees’ with elevated and decreased mean SBP of 20mmHg or more were 5.2% and 4.9% after 15min post-first dose vaccination, respectively. For the elevated SBP group (n=14), mean of SBP changes were 25mmHg (95%CI: 23-28mmHg) while the decreased SBP group (n=15) had a mean of SBP changes of 27mmHg (95CI%:23-30mmHg). Nevertheless, upon receipt of the second dose in the same cohort, an increase in the percentage of vaccinees (11%) had a decrease in SBP ≥20mmHg with mean SBP changes of 27mmHg (95% CI: 25-30mmHg) 15-minute post-vaccination, compared to 3.1% who had elevated SBP≥20mmHg (mean SBP changes: 26mmHg, 95%CI: 24-28mmHg). Nevertheless, we concluded that this was not a true SBP reduction. We noted that the pre-vaccination SBP was higher to begin with, upon the second dose as compared to the first dose.

**Table 2:** BP Trends, Mean Changes between Pre- and Post- BNT162b2 Vaccination for Both-Doses Analysis.

| Blood Pressure Trends | First dose (n=287) |  |  |  |  | Second dose (n=287) |  |  |  |  |
| --- | --- | --- | --- | --- | --- | --- | --- | --- | --- | --- |
|  | Frequency, n (%) | BP changes, Mean, mmHg | Standard Deviation, mmHg | Standard Error | 95% Confidence interval, mmHg | Frequency, n (%) | BP Changes Mean, mmHg | Standard Deviation, mmHg | Standard Error | 95% Confidence interval, mmHg |
| **Systolic** |  |  |  |  |  |  |  |  |  |  |
| *Elevated, | 15 (5.2) | 25 | 5 | 1.33 | 23 to 28 | 9 (3.1) | 26 | 3 | 1.08 | 24 to 28 |
| †Decreased | 14 (4.9) | 27 | 6 | 1.69 | 23 to 30 | 32 (11.1) | 27 | 7 | 1.20 | 25 to 30 |
| ‡No Change | 258 (89.9) | 0 | 9 | 1 | -1 to 1 | 246 (85.7) | 1 | 9 | 0.58 | -1 to 2 |
| **Diastolic** |  |  |  |  |  |  |  |  |  |  |
| *Elevated | 19 (6.6) | 15 | 5 | 1.10 | 13 to 17 | 26 (9.1) | 15 | 9 | 1.64 | 12 to 19 |
| †Decreased | 72 (25.1) | 15 | 6 | 0.68 | 14 to 17 | 36 (12.5) | 13 | 6 | 0.97 | 12 to 15 |
| ‡No Change | 196 (68.3) | 1 | 5 | 0.34 | 1 to 2 | 225 (78.4) | 0 | 5 | 0.2 | -1 to 1 |
| **Pulse Pressure** |  |  |  |  |  |  |  |  |  |  |
| Widened | 82 (28.6) | 17 | 8 | 0.81 | 16 to 19 | 37 (12.9) | 15 | 6 | 0.93 | 14 to 17 |
| Narrowed | 35 (12.2) | 15 | 5 | 0.77 | 13 to 16 | 62 (21.6) | 18 | 9 | 1.17 | 16 to 21 |
| No Change | 170 (59.2) | -1 | 5 | 0.38 | -1 to 1 | 188 (65.5) | 1 | 12 | 0.36 | 1 to 4 |

\*the SBP, DBP, and PP increased by $\geq 20$ mmHg, $\geq 10$ mmHg, and $\geq 10$ mmHg, respectively.
†the SBP, DBP, and PP decreased by $\geq 20$ mmHg, $\geq 10$ mmHg, and $\geq 10$ mmHg, respectively
‡the SBP, DBP, and PP were within the normal range.

Interestingly, 15 minutes after the first dose, one quarter of vaccinees (25.1%) had a drop of DBP ≥ 10mmHg compared to only 6.6% who were noted to have elevated DBP ≥ 10mmHg **(Table 2)**. In the group with decreased DBP, mean BP changes was 15mmHg (95% CI: 14-17mmHg), while changes in the elevated DBP group was 15mmHg (95% CI: 13-17mmHg). Upon receiving the second dose, the percentage of vaccinees with decreased DBP ≥10mmHg was also high (12.5%), compared to those who had elevated DBP ≥ 10mmHg (9.1%). DBP changes for the decreased group was 13mmHg (95%CI: 12-15mmHg) and 15mmHg for the elevated group (95% CI: 12-19mmHg). Interestingly, a higher proportion of the vaccinees had reduced DBP ≥10mmHg after receiving the first dose compared the second dose (25.1% vs 12.5%).

There were high numbers of vaccinees with widened PP (28.6%, n=82) compared to those who had narrowed PP (12.2%, n=35) after receiving the first dose of vaccine. Percentages of vaccinees with widened and narrowed PP were 28.6% (mean PP changes: 17mmHg, 95% CI: 16-19mmHg) and 12.2% (mean PP changes: 15mmHg, 95% CI: 13-16mmHg), respectively, comparing between pre- and 15min post-first dose of vaccination **(Table 2)**. However, upon receiving the second dose, there was a paradoxical observation in terms of PP trends compared to the first dose. After the second dose, percentage of vaccinees who had narrowed PP (21.6%, mean PP changes: 18mmHg, 95%CI: 16-21mmHg) almost doubled whilst percentage of vaccinees with widened PP reduced by almost half. (12.9%, mean PP changes: 15mmHg, 95% CI: 14-17mmHg).

### VACCINATION AND BLOOD PRESSURE IN BOTH-DOSES ANALYSIS

The analysis of both-doses’ data is shown in **Table 3**. After the vaccinees received their first dose, mean DBP was observed to decrease by 3mmHg, mean MAP was decreased by 2 mmHg, and mean PP was widened by 3mmHg, respectively, 15 minutes post-vaccination as compared to pre-vaccination. However, the mean SBP was unchanged for first dose. The BP analysis for second dose vaccination showed a reduction of mean SBP by 3mmHg, mean MAP decreased by 2mmHg, and a paradoxical trend of mean PP, which was narrowed by 3mmHg at 15 minutes after receiving it. Mean DBP remained the same for the second dose.

**Table 3:**
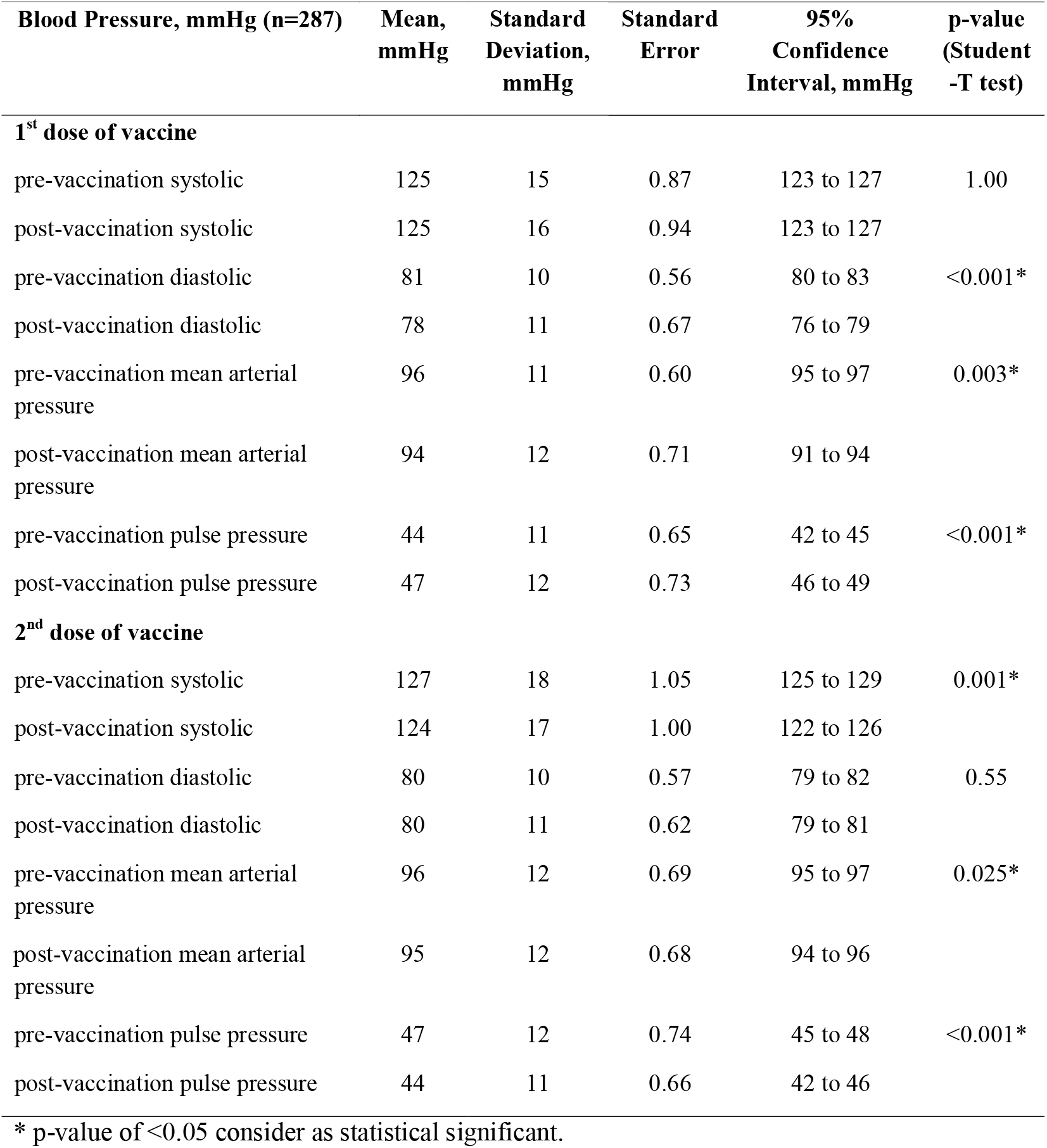
Comparison of BP Measurement Pre- and 15-minute Post- BNT162b2 Vaccination in Both-Doses Analysis.

## SUBGROUP ANALYSIS FOR FIRST DOSE VACCINATION

### TRENDS AND THE MEANS OF SBP, DBP, AND PP CHANGES BETWEEN PRE-AND 15-MINUTE POST-BNT162B2 VACCINATION IN FIRST-DOSE ANALYSIS

Data from a total of 431 vaccinees were included in this analysis. Detailed BP classifications for all vaccinees who received their first dose were shown in **Table S2**. We observed that there were a total of 5.3% and 5.6% of vaccinees who showed elevated and decreased in SBP mean differences, respectively (**Table 4**). The elevated SBP group had a mean SBP changes of 25mmHg (95%CI: 24-28mmHg) while the decreased SBP group had a mean SBP changes of 28mmHg (95%CI: 24-30)

**Table 4:** BP Trends between Pre-and 15-minutes Post- BNT162b2 Vaccination in First-Dose Analysis.

| **Blood Pressure Trends (n=431)** | **Frequency, n (%)** | **BP changes, Mean, mmHg** | **Standard deviation, mmHg** | **Standard Error** | **95% Confidence Interval, mmHg** |
| --- | --- | --- | --- | --- | --- |
| **Systolic** |  |  |  |  |  |
| *Elevated | 23 (5.3) | 25 | 5 | 1.08 | 24 to 28 |
| †Decreased | 24 (5.6) | 28 | 7 | 1.42 | 24 to 30 |
| ‡No Change | 384 (89.1) | 1 | 9 | 0.47 | -1 to 1 |
| **Diastolic** |  |  |  |  |  |
| *Elevated | 30 (7.0) | 14 | 4 | 0.79 | 13 to 16 |
| †Decreased | 108 (25.1) | 15 | 5 | 0.47 | 14 to 16 |
| ‡No Change | 293 (68.0) | 1 | 5 | 0.27 | 1 to 2 |
| **Pulse Pressure** |  |  |  |  |  |
| Widened | 104 (32.5) | 16 | 7 | 0.70 | 15 to 18 |
| Narrowed | 55 (12.8) | 14 | 5 | 0.69 | 14 to 16 |
| No Change | 262 (60.8) | 1 | 5 | 0.32 | -1 to 1 |

\*the SBP, DBP, and PP increased by $\geq 20\text{mmHg}$ , $\geq 10\text{mmHg}$ , and $\geq 10\text{mmHg}$ , respectively.
†the SBP, DBP, and PP decreased by $\geq 20\text{mmHg}$ , $\geq 10\text{mmHg}$ , and $\geq 10\text{mmHg}$ , respectively
‡the SBP, DBP, and PP were within the normal range.

The trend of DBP changes was like the both-doses analysis. There were 25.1% vaccinees with a decreased ≥10mmHg DBP compared to only 7% who had elevated DBP 15min post-vaccination. Mean DBP change for the decreased DBP group was 15mmHg (95%CI: 14-16mmHg) and for the elevated DBP group was 14mmHg (95%CI: 13-16mmHg).

There was an increase of vaccinee percentage observed to have widened PP ≥ 10mmHg, which was 32.5% (mean PP changes: 16mmHg, 95%CI: 15-18mmHg) compare to narrowed PP, which was 12.8% (mean PP changes: 14mmHg, 95CI%: 14-16mmHg).

### VACCINATION AND BLOOD PRESSURE

Database analysis for first-dose analysis is shown in **Table 5**. The results showed that mean DBP decreased by 4mmHg, mean MAP decreased by 3mmHg, and mean PP widened for 3mmHg, 15 minutes post-vaccination compared to pre-vaccination. Mean SBP has remained unchanged.

**Table 5:**
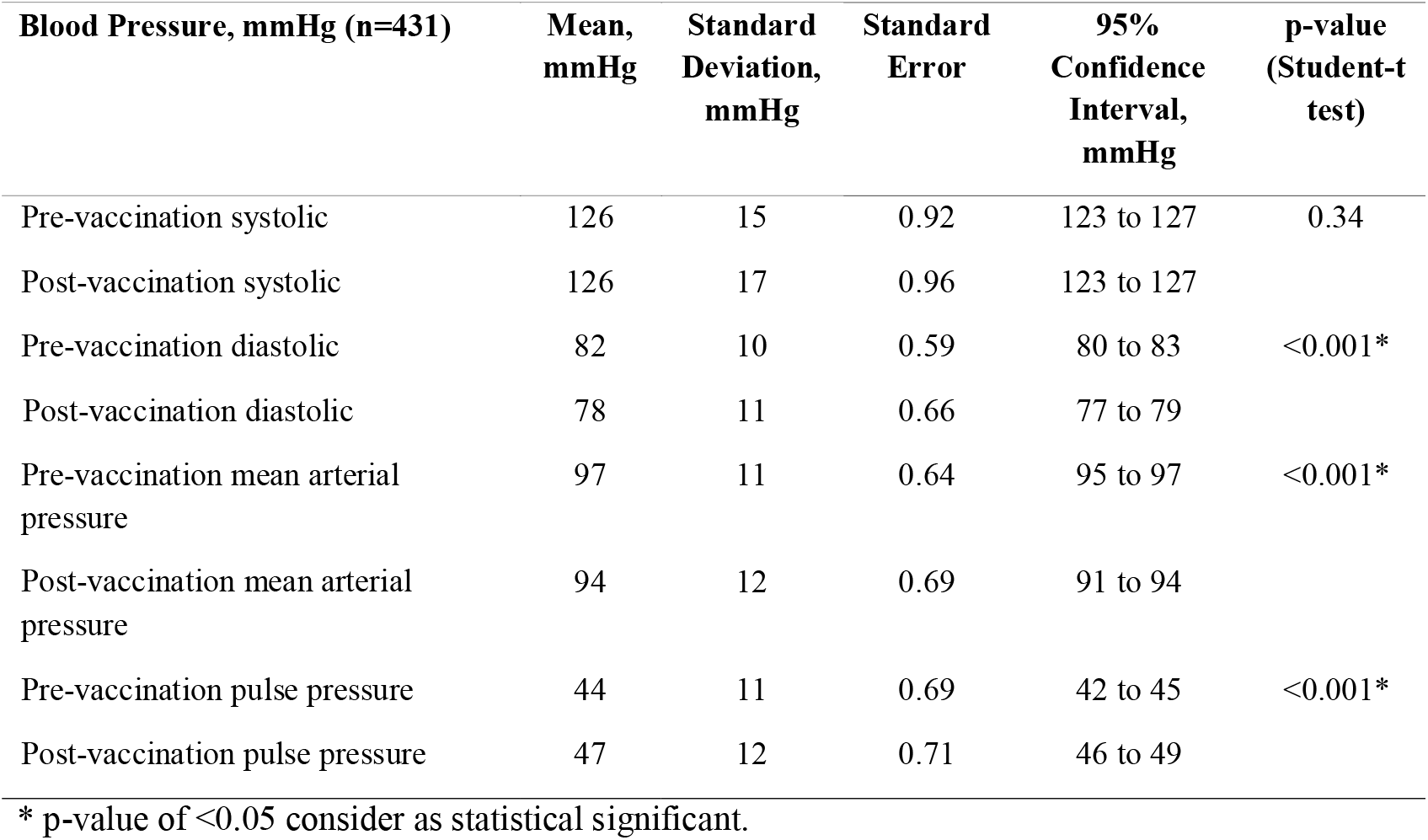
Comparison of BP Measurement Pre- and 15min Post- BNT162b2 Vaccination for First-Dose Analysis.

## DISCUSSION

Many are concerned about possible side effects from COVID-19 vaccination, (especially by the newly developed mRNA vaccines, including BNT162b2 from Pfizer) which was approved under EUA. One of the side effects of concern was the effect of the vaccine on BP. This concern seems valid as there was recently published case series on eight vaccinees who experienced elevated blood pressure after receiving the vaccine ^7^.

Based on our VSAS, a few noteworthy findings related to vaccinees’ BP after the BNT162b2 vaccination were found. Among all vaccinees who received the first dose of this vaccine, one quarter of vaccinees experienced a transient drop in DBP and one-third of them displayed widened PP. However, this phenomenon was not observed upon the second dose of vaccination. Interestingly, mean SBP remained unchanged after the first dose of the vaccine with minimal reduction after receiving the second dose. These findings of mean DBP reduction and mean PP widening after the first vaccination had not been reported elsewhere.

The findings of lower DBP and widened PP after the first dose of vaccine was unexpected. There are limited reports on transient effect compared to long term effect of DBP fluctuation. The study by Jaakko et. al. found that patients with pre-existing stages II and III hypertension whose DBP was below 90 mmHg had a higher risk of dying from cardiovascular disease, compared with patients with mean DBP in the range 90–109 mmHg^8^. The same study also concluded that lower DBP among patients with WHO stage I hypertension was not significantly associated with cardiovascular disease mortality. The study also found significant low DBP among pre-existing cardiac failure patients may increase the risk of cardiovascular death. The study also highlighted the increased risk of mortality for hypertensive patients above 50 years old with lower DBP. This study concluded that patients who had comorbid conditions such as neurovascular disease, ischaemic heart disease, and cancer with mean DBP below 90 mmHg were found to be associated with higher mortality than those with DBP in the range 90–109 mmHg. Study by Witteman et al. suggested that the decrease in DBP indicates vessel walls stiffening and parallels the degree of atherosclerosis progression^9^.

This transient BP fluctuation after vaccination, may present a hypothetical risk on the possibility of vaccinees developing neurovascular^10,11^ and cardiovascular adverse events, especially for those who have comorbid diseases^12-14^. No neuro- or cardiovascular events occurred in our active surveillance cohort, implying that the risk of these adverse events occurring due to the vaccination may be minimal. Our cohorts were rather young, and the majority were free of comorbidities. Furthermore, our cohorts were mainly HCWs and had easy access to the hospital facility. In the event of any major fluctuation of BP, they would have been immediately monitored at the on-site staff clinic or referred to the Emergency Department. Those who had comorbidities were also closely monitored after the vaccination.

On the other hand, our surveillance found that 10% of vaccinees were observed to have lower SBP after receiving the second dose of vaccine as compared to the first dose. However, this was not observed during the first dose of vaccination, which triggers the question whether it was directly related to the vaccine, or the psychological aspect of vaccinees. The vaccinees who may be anxious about the hype surrounding the higher AEFI risk after the second dose thus leading to higher pre-vaccination SBP^15^. This percentage of the increased pre-SBP may seem to be small, but its effect on stroke or cardiovascular events on healthy vaccinees are unclear^16^.

We also found about one-third of the vaccinees had wide pulse pressure after receiving the first dose of vaccine. The widened pulse pressure and lower DBP after the first dose of the vaccine may indicate the presence of vasodilation. This vasodilation may be caused by a mild anaphylaxis reaction possibly related to vaccines or their components. PP is the pulsatile component of repetitive continuous waves produced by BP that propagates along the arterial tree^17^. An increase of PP by 10 mmHg was found to increase the risk of a cardiovascular event, stroke, or overall mortality by 10– 20 percent. A few studies suggested that PP may be a better predictor for cardiovascular events among the middle-aged population ^18-20^. However, there was no report associating the transient changes of PP to vaccination adverse events.

The VSAS program was formed to monitor the safety of HCWs in HCTM. The surveillance actively monitors our centre’s vaccinee status and to look out for AEFI. The committee actively reports, manages, and monitors the vaccinees in the event of AEFI cases. Having a VSAS program ensures that potential AEFIs are promptly treated; this directly improves the confidence of vaccinees toward the COVID-19 vaccination program.

The strength of this study lies in its active surveillance design. The timing of BP measurements (pre- and 15-minute’s post-vaccination) was in the acceptable time frame to directly examine vaccine-related adverse events. The limitation of this surveillance was that there were many original HCWs who rescheduled their second dose of vaccination due to unavoidable logistic reasons.

## CONCLUSION

Our surveillance concluded that the BNT162b2 vaccination was found to be generally tolerated for participants vaccinated in our centre. Nevertheless, there might be a hypothetical neuro- and cardiovascular risk after the first dose, which may relate to DBP lowering and widened PP, especially for those with comorbidities. Our VSAS findings may provide a guide to policymakers for future roll-out of mass vaccination programs especially for vaccinees with comorbidities.

## Supporting information

D:\My research and publication\Original Articles\Vaccine COVid19 2021 AEFI study\BP study databased and manuscript\Manuscript\MedRxiv\Data Supplement.

## Data Availability

All vaccine-related data that were collected were kept in a secured location and only accessible to authorised personnel. The data that support the findings of this study are available from Toh Leong Tan. However, restrictions apply to the availability of these data, which were used under license for the current study, and so are not publicly available. Data are, however, available from the authors upon reasonable request and with permission of Toh Leong Tan.

## ACKNOWLEDGEMENTS

We thank colleagues who provided equipment and support materials for this study: all staff from The Department of Emergency Medicine, especially Dr. Faizal Amri, the head of the department, and Mr. Bala Krisnian, the head of assistance medical officer, the Infection Control Unit, and Pharmacy Department, HCTM. We would also like to thank all members of the HCTM Vaccine Committee and Prof. Dr. Mohd Shahrir Mohamed Said for their support. We would like to thank Assoc. Prof. Dr. Hui-min Neoh from UKM Medical Molecular Biology Institute for proofreading the manuscript. We would like to thank Prof. Dr. Simon Finfer from The George Institute for Global Health for the improvement of the manuscript.

## AUTHORS’ INFORMATION

From Department of Emergency Medicine, Faculty of Medicine, Universiti Kebangsaan Malaysia (TLT, ZCM); Department of Medical Microbiology and Immunology, Faculty of Medicine, Universiti Kebangsaan Malaysia (SA); The Pharmacy Department, Hospital Canselor Tuanku Muhriz, Universiti Kebangsaan Malaysia (MHPT); Quality Department, Hospital Canselor Tuanku Muhriz, Universiti Kebangsaan Malaysia (AR); and Department of Surgery, Faculty of Medicine, Universiti Kebangsaan Malaysia (RJ)

## CONTRIBUTIONS

T.L.T. and Z.C.M. initiated the conceptualisation of the study. T.L.T., S.A., Z.C.M. and M.H.P.T. wrote the methodology. T.L.T. and Z.C.M. handled the software. T.L.T., S.A., Z.C.M. and M.H.P.T. validated the data. T.L.T. and Z.C.M. did the formal analysis. T.L.T., S.A., Z.C.M., M.H.P.T., A.R. and R.J. performed the investigation. Resources were secure by T.L.T., S.A., Z.C.M., M.H.P.T., A.R. and R.J. Data curation were done by T.L.T. and Z.C.M. All the following: T.L.T., Z.C.M. and M.H.P.T. did the writing and prepared the draft. Write, review & editing were done by T.L.T., S.A., Z.C.M., M.H.P.T., A.R. and R.J. Visualisations were prepared by T.L.T. Project supervision, T.L.T.; Project administration run by T.L.T., S.A. and R.J. Funding Acquisition by T.L.T. All tables and figures were prepared by T.L.T. All authors have reviewed and agreed to the published version of the manuscript.

## ADDITIONAL INFORMATION

### Publisher’s note

Springer Nature remains neutral with regard to jurisdictional claims in published maps and institutional affiliations.

