## Supplementary material for "BNT162b2 COVID-19 VACCINATION AND ITS EFFECT ON BLOOD PRESSURE": D:\My research and publication\Original Articles\Vaccine COVid19 2021 AEFI study\BP study databased and manuscript\Manuscript\MedRxiv\Data Supplement.

**CONTENT**

| **List** | **Content** | **Page** |
| --- | --- | --- |
| **Table S1** | Detail Demography of the Vaccinees Who Had a History of Hypertension under Treatment in Both-Group Analysis | 3 |
| **Table S2** | The Blood Pressure Characteristics for All the Vaccinees in First-Dose Analysis. | 4 |
| **Figure S1** | Study population for BNT162b2 vaccines. | 5 |

**THE BNT162b2 COVID-19 VACCINATION AND THE EFFECT ON BLOOD PRESSURE**

**Table S1. Detail Demography of the Vaccinees Who Had a History of Hypertension under Treatment in Both-Group Analysis**

| **Age, years** | **Gender** | **First Dose of Vaccine Blood Pressure, mmHg** | | | | **Second Dose of Vaccine Blood Pressure, mmHg** | | | |
| --- | --- | --- | --- | --- | --- | --- | --- | --- | --- |
|  |  | **Pre-vaccination SBP** | **Pre-vaccination DBP** | **Post-vaccination SBP** | **Post-vaccination DBP** | **Pre-vaccination SBP** | **Pre-vaccination DBP** | **Post-vaccination SBP** | **Post-vaccination DBP** |
| 56-60 | Male | 187 | 78 | 152 | 54 | 164 | 78 | 164 | 77 |
| 46-50 | Male | 151 | 94 | 151 | 96 | 172 | 99 | 174 | 98 |
| 46-50 | Female | 140 | 71 | 150 | 93 | 139 | 93 | 133 | 96 |
| 41-45 | Female | 159 | 91 | 156 | 95 | 114 | 73 | 124 | 79 |
| 56-60 | Female | 155 | 87 | 156 | 85 | 145 | 83 | 145 | 89 |
| 41-45 | Female | 145 | 91 | 149 | 84 | 136 | 81 | 124 | 77 |
| 41-45 | Female | 108 | 79 | 103 | 71 | 140 | 79 | 144 | 105 |
| 41-45 | Female | 134 | 88 | 130 | 86 | 139 | 90 | 134 | 91 |

**Table S2. The Blood Pressure Characteristics for All the Vaccinees in First-Dose Analysis.**

| **Systolic Blood Pressure, mmHg (n=431)** | **Frequency, n (%)** |
| --- | --- |
| Pre-vaccination |  |
| Normal: < 120 | 150 (34.8) |
| Prehypertension: 120-139 | 206 (47.8) |
| Stage 1:140 -159 | 60 (13.9) |
| Stage 2: ≥160 | 15 (3.5) |
| 15min Post-vaccination |  |
| Normal < 120 | 156 (36.2) |
| Prehypertension 120-139 | 189 (43.9) |
| Stage 1:140 -159 | 75 (17.4) |
| Stage 2 : ≥160 | 11 (2.6) |
| **Diastolic Blood Pressure, mmHg** |  |
| Pre vaccination |  |
| Normal: <80 | 177 (41.1) |
| Prehypertension: 80-90 | 168 (39.0) |
| Stage 1: 90- 99 | 67 (15.5) |
| Stage 2 : ≥100 | 19 (4.4) |
| 15min Post-vaccination |  |
| Normal: <80 | 234 (54.3) |
| Prehypertension: 80-90 | 128 (29.7) |
| Stage 1: 90- 99 | 54 (12.5) |
| Stage 2 : ≥100 | 15 (3.5) |

**HCTM COVID-19 vaccination safety surveillance database:**

15^th^ -16^th^ March 2021(first dose) and

5^th^ -6^th^ April 2021 (second dose)

**Data fulfil the inclusion**:

*Both doses completed (n=288)

First dose only (n=443)

**Data includes**:

Age, gender, blood pressure pre-vaccination; blood pressure 15min post-vaccination and Adverse Events

**Exclusion:**

Missing BP data for both dose analysis: n= 1 (0.3%)

Missing BP data for first dose subgroup analysis: n=12 (2.8%)

**Data Eligible for Analysis**

Both-doses analysis (n=287)

**Subgroup analysis**

First-dose analysis (n=431)

**Figure S1. Study population for BNT162b2 vaccines.**

*Vaccinees had rescheduled their second dose to another day due to logistic reasons (n=143). They were excluded from both doses analysis.
